# Regulatory Variants on the Leukocyte Immunoglobulin-Like Receptor Gene Cluster are Associated with Crohn’s Disease and Interact with Regulatory Variants for *TAP2*

**DOI:** 10.1101/2023.03.28.23287842

**Authors:** Kwangwoo Kim, Shin Ju Oh, Junho Lee, Ayeong Kwon, Chae-Yeon Yu, Sangsoo Kim, Chang Hwan Choi, Sang-Bum Kang, Tae Oh Kim, Dong Il Park, Chang Kyun Lee

## Abstract

**Background and Aims:** Crohn’s disease (CD) has a complex polygenic etiology with high heritability. We keep putting an effort to identify novel variants associated with susceptibility to CD through a genome-wide association study (GWAS) in large Korean populations.

**Methods:** Genome-wide variant data from 902 Korean patients with CD and 72,179 controls were used to assess the genetic associations in a meta-analysis with previous Korean GWAS results from 1,621 patients with CD and 4,419 controls. Epistatic interactions between CD-risk variants of interest were tested using a multivariate logistic regression model with an interaction term.

**Results:** We identified two novel genetic associations with the risk of CD near *ZBTB38* and within the leukocyte immunoglobulin-like receptor (LILR) gene cluster (*P*<5×10^−8^), with highly consistent effect sizes between the two independent Korean cohorts. CD-risk variants in the LILR locus are known quantitative trait loci (QTL) for multiple LILR genes, of which *LILRB2* directly interacts with various ligands including MHC class I molecules. The LILR lead variant exhibited a significant epistatic interaction with CD-associated regulatory variants for *TAP2* involved in the antigen presentation of MHC class I molecules (*P*=4.11×10^−4^), showing higher CD-risk effects of the *TAP2* variant in individuals carrying more risk alleles of the LILR lead variant (OR=0.941, *P*=0.686 in non-carriers; OR=1.45, *P*=2.51×10^−4^ in single-copy carriers; OR=2.38, *P*=2.76×10^−6^ in two-copy carriers).

**Conclusions:** This study demonstrated that genetic variants at two novel susceptibility loci and the epistatic interaction between variants in LILR and *TAP2* loci confer risk of CD.

## 1. Introduction

Crohn’s disease (CD) is a complex inflammatory bowel disease with strong genetic components. The risk of developing CD is significantly higher in individuals with a family history of the disease compared to the general population.^1^ Twin studies estimated the heritability of CD to be 75%, showing that the disease concordance rate for CD is much higher in monozygotic twins than in dizygotic twins.^2^

Genome-wide association studies (GWASs) have been highly productive in identifying variant-level genetic components associated with susceptibility to CD, including *IL23R, NOD2*, and others.^3^ To date, the number of reported CD loci is over 240 that have opened up new avenues to understand the underlying mechanisms of disease development and to identify potentially effective drug targets for CD. However, the highly polygenic genetic architecture of CD resulted in a large number of CD-risk variants remaining hidden in the human genomes, leading to considerably large missing heritability. GWAS variants, including reported CD loci, explain a small fraction of the heritability of CD,^2^ suggesting that a continuous effort to identify CD-risk variants is necessary to better understand the dysregulation of the gastrointestinal immune system in patients with CD. Indeed, a Bayesian simulation approach that inferred the genetic architecture of common complex disorders suggested the presence of hundreds of common causal variants with weak to modest effect sizes as well as multiple rare variants with strong effects.^4^ In addition, the non-additive epistatic effects among independently inherited genetic variants are largely unknown; however, these are expected to substantially explain a proportion of the missing heritability.^5^

Moreover, GWASs in non-European individuals have been relatively infrequent.^6^ Considering the ethnicity-associated genetic heterogeneity and the limited ethnic transferability of polygenic risk scores, it is crucial to conduct more genetic studies in relatively understudied populations, including the Korean population. In recent decades, the incidence and prevalence of inflammatory bowel disease has been steadily increasing in Korea.^7^

In this study, we generated genome-wide variant data from a large Korean population and performed a meta-analysis with existing Korean GWAS association summary statistics to identify robust novel association signals and gene-gene interactions to expand the knowledge of the genetic etiology of CD.

## 2. Materials and Methods

### 2.1. GWAS subjects

Our case-control GWAS newly analyzed the individual-level genotype data of 902 patients with CD and 72,179 healthy controls, with a subsequent meta-analysis with previous GWAS association statistics^8^ estimated from 1,621 patients with CD and 4,419 controls. Among the newly analyzed subjects, patients with CD were recruited from Kyung Hee University Hospital (KHUH; n=253 from a subset of a multi-omics study cohort with a ClinicalTrials.gov identifier NCT03589183) and 15 tertiary hospitals (n=649 from a retrospective multicenter CD cohort from the IMPACT study^9^). All patients met the diagnostic criteria of CD based on clinical, serological, and radiological results as well as endoscopy and histology results.^10^ Patients with indeterminate colitis were excluded. All study participants were aged ≥16 years at study enrollment and provided peripheral blood samples for genomic DNA extraction. Controls were recruited through the Korean Genome Epidemiology Study (KoGES), providing genomic DNAs to the National Biobank of Korea.

The study was conducted in accordance with the ethical principles of the Declaration of Helsinki and was approved by the Institutional Review Board of the participating hospitals (KHUH 2018-03-006-018, KHUH 2021-04-082, KBSMC 2016-07-029). All participants provided written informed consent for participation in the study.

### 2.2. Genetic variant data

Genome-wide variant data were generated using a custom Affymetrix array named KoreanChip^11^ developed by the Korea National Institute of Health. The array comprised over 800,000 variants yielding high coverage of genomic variants with moderate to high imputation quality, particularly for Korean-specific, functional, and low-frequency variants.^11^ After a general quality control procedure for genotyping data (**Supplementary Table 1**), whole-genome imputation for 902 cases and 72,179 controls was performed based on the reference panel of the 1000 Genomes Project (1KGP) phase 3 through the Michigan Imputation Server^12^ using Eagle2 and Minimac4. A total of 8,950,452 unique variants with minor allele frequencies (MAFs) ≥ 0.5%, Hardy-Weinberg equilibrium (HWE; *P*-values for HWE ≥ 10^−6^ in controls and 10^−10^ in cases), and imputation accuracy values R2 ≥ 0.3 were included in all subsequent statistical analyses. HLA imputation was performed separately by the Michigan Imputation Server using Eagle2 and Minimac4 based on the multi-ethnic HLA reference panel^13^.

### 2.3. Statistical tests

#### 2.3.1. Single-marker association

A single-marker association for CD in our study population was assessed using multivariate logistic regression adjusted for the top five principal components and sex to calculate an odds ratio (OR) and its 95% confidence interval (CI) for each variant. We defined the boundaries of each susceptibility locus by merging the positions of the CD-associated variants in the 250 kb flanking regions of each variant so that the susceptibility loci were at least 500 kb apart from each other. In addition, we visually inspected the regional association plots for each locus to confirm the independence of the CD associations.

#### 2.3.2. Meta-analysis with previous Korean GWAS results

A previous genome-wide association summary statistics dataset for Korean CD was retrieved from Jung *et al*.^8^ This dataset included 5,885,366 variants from an independent Korean cohort consisting of 1,621 CD cases and 4,419 controls, after excluding 307 variants with mismatched allele codes from dbSNP v150. A GWAS meta-analysis of our newly generated GWAS results with the previous GWAS results was performed by the METAL program^14^ using an inverse-variance fixed-effects model after genomic control for each individual dataset, bringing the final total sample size to 79,121 (n = 2,523 CD cases and 76,598 controls). The extended major histocompatibility complex (MHC) region in a high linkage disequilibrium (LD) was assumed to span from 25 to 36 Mb on chromosome 6.

To identify a credible set of the most likely causal variants associated with CD in each susceptibility locus, a Bayesian fine-mapping approach using FINEMAP v1.4 was applied to estimate the posterior probability (PP) for each variant to be causal based on the meta-analysis results and the in-sample LD from the newly analyzed GWAS samples.^15^ A 95% credible set has the minimum number of the best causal candidate variants whose cumulative sum of PPs is greater than 0.95.

#### 2.3.3. Estimation of heritability from GWAS variants

A heritability of CD was calculated from all genome-wide variants listed in our meta-analysis by using LD score regression (LDSC)^16^, based on the genetic architecture of the 1KGP East Asians. The latest prevalence of CD was 0.00030 in the Korean population^7^; therefore, this value was used to compute heritability values on the liability scale.

#### 2.3.4. Epistatic interaction test

Epistatic interaction tests were performed to examine whether a joint effect between two genetic variants of interest deviated significantly from a multiplicative joint effect. For this, a multivariate logistic regression model with an interaction term between the two variants was employed using the newly generated GWAS dataset, with adjustments for the top five principal components and sex. Specifically, we evaluated the epistatic interaction between a novel CD-risk lead variant of interest and potentially functional CD-risk variants in genes related to the MHC class I-mediated antigen presentation pathway. Pathway gene members (n = 28) were retrieved from the gene ontology term ‘antigen processing and presentation of peptide antigens via MHC class I’ (GO: 0002474) using the msigdbr package^17^. Functional variants included protein-altering variants and expression quantitative trait loci (eQTL). To collect the most relevant eQTL variants, we extracted only the lead eQTL variants for each gene in the pathway based on the previous eQTL results^18^ from six different immune cell types of Japanese individuals that share similar genetic characteristics with Koreans in terms of allelic frequency and LD.

## 3. Results

### 3.1. Identification of three novel susceptibility loci for CD

We performed a meta-analysis of the genome-wide association results of the newly generated CD GWAS data and previously published GWAS data^8^, providing the disease association statistics for 9,044,589 autosomal variants in 2,523 CD cases and 76,598 controls. The genomic-control inflation factor (λ_GC_) was 1.038 and the LDSC intercept was 0.974, indicating little evidence of systemic bias and population stratification in the study cohort. A principal component analysis also supported a highly consistent genetic background between the cases and controls (**Supplementary Figure 1**).

There were 1,227 variants surpassing the genome-wide significance threshold in 28 non-MHC loci (**Figure 1**). Of these, there were two novel association signals at rs255774 within the leukocyte immunoglobulin-like receptor (LILR) gene cluster (OR = 1.32, 95%CI = 1.22 to 1.42, *P*_meta_ = 1.71 × 10^−12^) and at rs6802753 near *ZBTB38* (OR = 1.28, 95%CI = 1.18 to 1.38, *P*_meta_ = 4.45 × 10^−10^). The lead variant in each novel susceptibility locus showed a modest effect size, which was highly consistent between the two Korean datasets (**Table 1**). In addition, the associations of 26 reported non-HLA loci, including *TNFSF15-TNFSF8, DUSP5, GPR35*, and *NOD2*, were replicated in the Korean population (**Supplementary Table 2**). The liability-scale narrow-sense heritability *h*^*2*^ estimated from the GWAS meta-analysis variants was 19.1%, of which approximately 3.0% accounted for the two novel loci.

**Figure 1.**
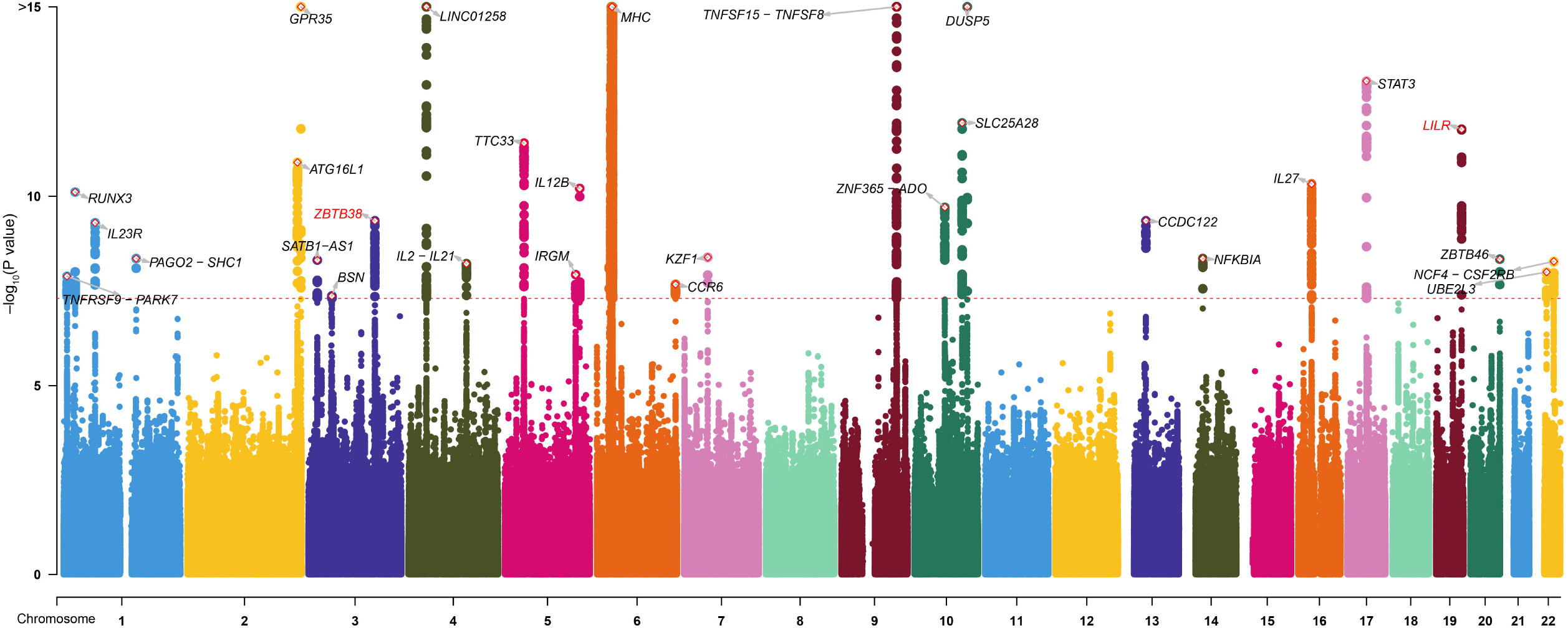
Manhattan plot for the genome-wide association meta-analysis of Crohn’s disease in the Korean population. Novel and known susceptibility loci are labeled in red and black, respectively.

**Table 1.** Genetic associations of two novel susceptibility loci for Crohn’s disease in Korean populations.

| Variant | Chr:Position (hg19) | EA/NEA | Korean dataset #1 (new) |  | Korean dataset #2 (Jung *et al.*) |  | Meta-analysis |  |  |
| --- | --- | --- | --- | --- | --- | --- | --- | --- | --- |
|  |  |  | OR (95% CI) | *P* | OR (95% CI) | *P* | EAF | OR (95% CI) | *P*<sub>meta</sub> |
| rs255774 | 19:54723355 | G/A | 1.34 (1.20–1.50) | 1.94×10<sup>-7</sup> | 1.30 (1.18–1.43) | 1.92×10<sup>-7</sup> | 0.456 | 1.32 (1.22–1.42) | 1.71×10<sup>-12</sup> |
| rs6802753 | 3:141145473 | C/T | 1.39 (1.24–1.55) | 5.09×10<sup>-9</sup> | 1.19 (1.08–1.31) | 6.29×10<sup>-4</sup> | 0.253 | 1.28 (1.18–1.38) | 4.45×10<sup>-10</sup> |

The genetic effect sizes between the two datasets were not statistically different (*P* values for heterogeneity > 0.05).
Chr, Chromosome; EA, effect allele; NEA, non-effect allele; OR, odds ratio; CI, confidence interval; EAF, effect allele frequency.

### 3.2. Functional annotation of CD-risk variants

#### 3.2.1. LILR gene cluster locus

We identified multiple variants associated with CD in the LILR gene cluster region (**Figure 2**A). A Bayesian fine-mapping analysis suggested a credible variant set in which the listed variants had a relatively high posterior probability of being causal (**Supplementary Table 3**). These variants (n = 5) with over 95% cumulative probability of being causal were all located in non-coding elements and have previously been associated with the expression or alternative splicing of multiple genes, including *LILRB2, LILRB3, LILRB4*, and *LILRA6*, in whole blood according to GTEx Analysis Release V8 (dbGaP Accession phs000424.v8.p2).

**Figure 2.**
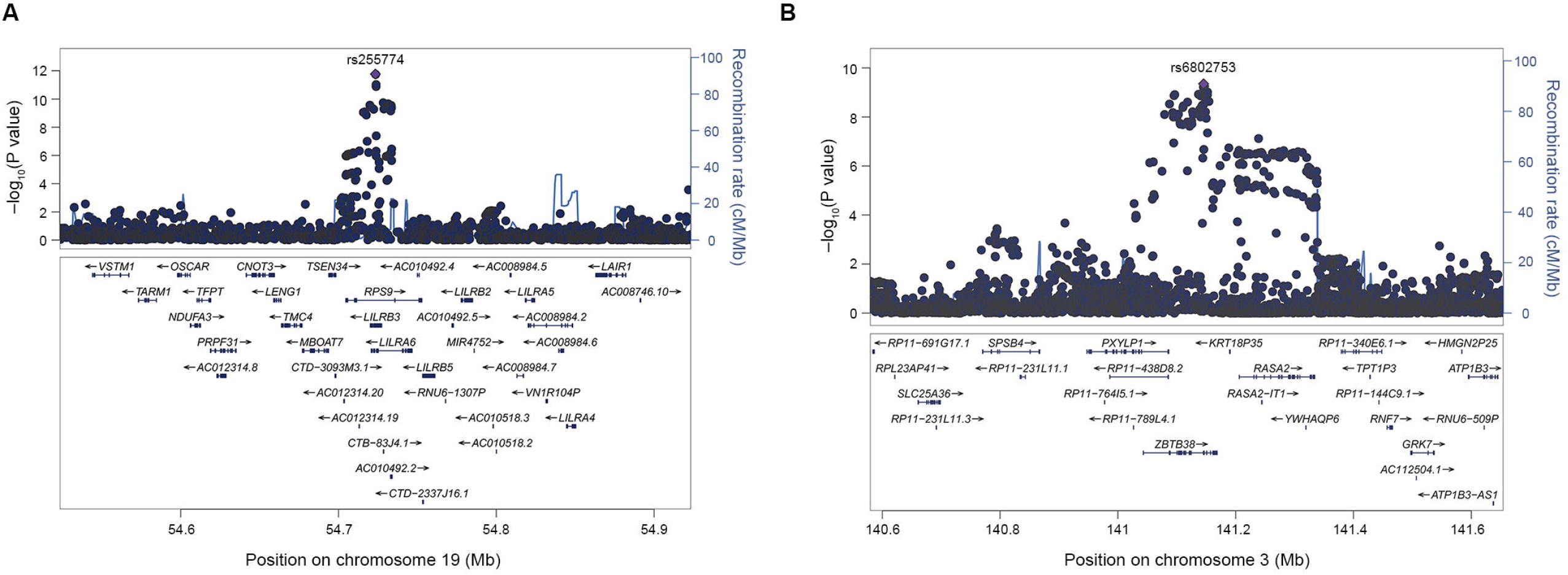
Regional association plots of the novel susceptibility loci for Crohn’s disease. **(A)** *ZBTB38* locus **(B)** leukocyte immunoglobulin-like receptor gene cluster locus.

The lead variant rs255774 was detected as a splicing QTL (sQTL)^19^ associated with the variation in intron splicing efficiency of *LILRB2* in whole blood (GTEx splicing phenotype number: chr19:54218413:54276264:clu21769). The intron splicing associated with the CD-risk allele *C* of rs255774 results in the exclusion of all three immunoreceptor tyrosine-based inhibitory motifs (ITIMs) in the cytoplasmic tail of LILRB2 proteins that interact directly with MHC class I molecules, even when these are present on the same cells.^20^ Therefore, as the ITIMs inhibit MHC class I-mediated activating signals, it is plausible that the CD-risk variant downregulates an inhibitory pathway of the MHC class I-mediated immune response.

Various approaches have long demonstrated the importance of antigen presentation by MHC class I molecules in the pathogenesis of many inflammatory diseases, including CD. In particular, the genetic variants in the MHC region explained a large fraction of the known genetic predisposition to CD in multiple ancestries, and an HLA association fine-mapping study clearly pinpointed the genetic contribution of MHC class I genes, such as *HLA-B* and *HLA-C*, to the development of CD.^21^

#### 3.2.2. The *ZBTB38* locus

The *ZBTB38* locus has been well documented owing to the pleiotropic effects of its genetic variants on various human traits, including height.^22^ Notably, the lead variant rs6802753 and its LD proxies (*r*^2^ > 0.6) with genome-wide association significance were limited to *ZBTB38* (**Figure 2**B). Due to the high LD in the locus, it is challenging to statistically narrow the list of these variants down to a small number of putative causal candidate variants. There were 39 variants in the 95% credible set; however, none of the variants had PPs of over 0.1 (**Supplementary Table 4**).

Some of the variants in the 95% credible set of the *ZBTB38* locus were located with active chromatin states^23^, and one of the proxies (rs6440008 with *P*_meta_=2.31×10^−9^) was previously associated with the expression level of *ZBTB38* in lymphoblastoid cell lines.^24^ In addition, although none of proxies in a strong LD with the lead variant resided in protein-coding elements, there was a relatively weakly correlated nonsynonymous proxy, rs62282002 (*r*^*2*^=0.61), on *ZBTB38* with a suggestive CD association (*P*_meta_=4.75×10^−6^), which was previously reported to be associated with serum total protein levels^25^ and idiopathic short statures^26^.

### 3.3. Identification of the epistatic interaction between the LILR variant rs255774 and *TAP2*-regulatory variants

Several LILRs, including *LILRB2*, are known to recognize a broad range of ligands, including MHC class I molecules, and contribute to the regulation of the MHC class I-mediated immune response.^27^ In this study, we aimed to identify the epistatic interaction between the lead CD-risk variant rs255774 in the LILR locus and CD-risk functional variants (lead eQTL or HLA variants) in the genes involved in the MHC class I-mediated antigen presentation using the newly generated GWAS dataset. There were 28 genes listed in the GO term “antigen processing and presentation of peptide antigens via MHC class I” (GO:0002474). Among these, 11 genes within the extended MHC region showed significant CD associations in our GWAS (**Supplementary Table 5**). Of the 11 genes, nine had significant eQTLs in various immune cell types of individuals with East Asian ancestry (lead_eQTLs:gene:tissue pairs = 29; **Supplementary Table 6**), and *HLA-B* showed significant CD associations at the level of HLA classical alleles and amino acid residues (**Supplementary Table 7**).

The multivariate logistic regression with an interaction term between the LILR variant rs255774 and putatively functional CD-risk variants in the MHC class I pathway revealed that the LILR variant rs255774 interacts with a lead eQTL for *TAP2* (*P*_interaction_ = 4.11 × 10^−4^) among the functional variants (**Supplementary Table 6**). Specifically, two CD-risk alleles in LILR and *TAP2* synergistically conferred risk of CD (OR_intereaction_=1.496). For example, when the study subjects were stratified according to the number of the CD-risk alleles of the LILR variant rs255774, the effect sizes of the CD-risk allele of *TAP2*-regulatory variant rs9276627 increased in individuals carrying more CD-risk LILR alleles (**Table 2**) and vice versa (**Supplementary Table 8**). In addition, the effect allele frequencies of rs9276627 were highly consistent among the rs255774-stratified control groups in contrast to the case groups, which indicates that the identified interaction follows the assumption of independence between two variants in the general population. Indeed, TAP2 transports peptide fragments from cytosol to the endoplasmic reticulum lumen for antigen processing and presentation mediated by MHC class I molecules, and the CD-risk variant of the *TAP2* eQTL rs9276627 is associated with increased expression of *TAP2*.

**Table 2.** Synergistic interaction effect of two quantitative trait loci for *TAP2* and LILR genes on susceptibility to Crohn’s disease.

| No. of CD-risk alleles of<br>the LILR variant rs255774 | No. of cases/controls | CD association of the *TAP2* lead eQTL rs9276627 |  |  |
| --- | --- | --- | --- | --- |
|  |  | EAF (cases/control) | OR (95% CI) | *P* value |
| 0 | 167/19,472 | 0.841/0.850 | 0.941 (0.700-1.264) | 0.686 |
| 1 | 506/36,637 | 0.891/0.850 | 1.452 (1.189-1.773) | 2.51x10<sup>-4</sup> |
| 2 | 229/16,070 | 0.930/0.850 | 2.376 (1.655-3.412) | 2.76x10<sup>-6</sup> |

Effect and non-effect alleles of rs9276627 are C and T, respectively.
CD, Crohn's disease; eQTL, expression quantitative trait locus; EAF, effect allele frequency; OR, odds ratio; CI, confidence interval.

## Discussion

There is the growing list of susceptibility loci for CD mostly through GWASs, but a substantial proportion of CD susceptibility loci remains unidentified.^2^ Our large-scale genome analysis, which took advantage of the Korean-specific high-coverage GWAS array, the so-called KoreanChip, as well as a meta-analysis using another ancestry-matched GWAS dataset, successfully identified and characterized two dependable CD susceptibility loci with highly consistent effect sizes in two independent case-control datasets of Korean individuals.

The novel loci identified in this study further contributed to 3.0% of the SNP-based heritability in the Korean cohort. The Bayesian inference of posterior inclusion probability was used to suggest the list of the most likely causal variants in the two novel loci. This finding supports the crucial role of non-coding variants in the pathogenesis of complex polygenic disorders. In particular, we identified five variants in a credible set, including potentially regulatory variants in the LILR locus. Of these, the two most significant variants explained >0.75 of the cumulative sum of PP. Notably, the lead CD-risk variant was an sQTL for *LILRB2* and an eQTL for multiple genes, including *LILRB3, LILRB4, LILRA6*, and *RPS9*.

Regarding the LILR locus, the CD-relevant effector genes were not precisely determined, given the multiple target genes of CD-associated QTLs and the limited knowledge about the specific functions of each candidate gene in the gastrointestinal immune system. Experimental approaches and genetic analyses using different ancestries are necessary to identify disease-relevant genes. Nevertheless, we suspect that *LILRB2*, interacting with MHC class I molecules, may confer risk of CD via a synergistic gene-gene interaction with *TAP2* because risk variants on two genes in the same pathway may have a greater chance of interacting with each other in a biological sense. We identified the presence of an additional CD-risk effect on individuals with both the CD-risk QTL alleles for LILR genes and *TAP2* in a dose-dependent manner.

TAP2 is a member of the ATP-binding cassette transporter family and is involved in transferring cytosolic peptide fragments into the endoplasmic reticulum lumen, after which the fragments are loaded on the MHC class I molecules before being presented on the cell surface.^28^ The expression of *TAP2* is upregulated to facilitate the MHC class I-mediated antigen presentation in various cell types including macrophage and dendritic cells.^29^ In our study, the results indicate that the risk allele *C* of the CD-associated *TAP2* eQTL rs9276627 is associated with increased levels of *TAP2*. Consistently, elevated expression levels of *TAP2* was observed in patients with CD through a colon transcriptome analysis.^30^ Therefore, the risk alleles of both the sQTL for *LILRB2* and the eQTL for *TAP2* appear to be associated with the upregulation of the MHC class I-mediated immune response.

Another novel CD association at rs6802753 on 3q23 suggests that *ZBTB38* is the sole candidate causal gene. *ZBTB38* encodes “Zinc-finger and BTB Domain Containing 38” that regulates transcription while binding to methylated DNA elements.^31^ GWASs have demonstrated that variants in *ZBTB38* have pleiotropic effects on several human traits, including height,^32^ cancer,^33^ and prion disease^34^. The CD-risk variant rs6440008 in a high LD with the lead variant rs6802753 is a known eQTL whose CD-risk allele *C* is associated with the upregulation of *ZBTB38*.

Nevertheless, the exact physiological and immunological role of *ZBTB38* have been understudied. Through immunological approaches, *ZBTB38* was reported to be hypomethylated and transcriptionally upregulated in adaptive NK cells.^35^ Similarly, the murine homolog gene *Zbtb38* was highly expressed in a murine model of rheumatoid arthritis due to its hypomethylated promoter and interfered with an anti-inflammatory pathway by transcriptionally repressing *IL1r2* in B cells.^36^ In non-immune cells, the knockdown of *Zbtb38* resulted in apoptosis.^37^ In addition, a cancer study showed that increased expression of *ZBTB38* in human bladder cancer induced a more aggressive phenotype with enhanced migration and invasive growth through modulation of the Wnt/β-catenin signaling pathway.^38^

In summary, we identified two novel susceptibility loci for CD and an epistatic interaction effect between CD-risk variants on risk of disease. Given the novel putative causal genes and variants identified, this GWAS provides novel insights into the underlying pathogenesis of CD involving the dysregulation of both LILR genes in the MHC class I-mediated antigen presentation pathway and *ZBTB38*.

## Data Availability

All data produced in the present study are available upon reasonable request to the authors.

## Acknowledgements

We would like to thank the following individuals and group for their contributions to this work: the IMPACT study group for the contribution to genotyping for patients with Crohn’s disease; study coordinators in all participating hospitals for their help in recruiting subjects; professors Ho-Su Lee and Byong Duk Ye at the University of Ulsan College of Medicine for providing the GWAS summary statistics from their previous CD GWAS.

## Author Contributions

KK and CKL designed the study. KK, SJO, JHL, and CKL wrote the manuscript. SJO, SK, CHC, SBK, TOK, DIP, HJK, and CKL recruited/characterized patients with CD, obtained DNA samples, and/or generated GWAS data. KK, SJO, HJK, and CKL obtained genetic data from the KoGES under the permission of Centers for Disease Control of Korea. KK, JL, AK, and CYY analyzed genetic data. All authors participated in critical revision of the manuscript and approved the final version.

## Data Availability

The data used and analyzed for this study are available from the corresponding author upon reasonable request.

