## Supplementary materials for "Regulatory Variants on the Leukocyte Immunoglobulin-Like Receptor Gene Cluster are Associated with Crohn’s Disease and Interact with Regulatory Variants for *TAP2*"

*Kwangwoo Kim et al.*

### Supplementary Table 1. Quality control (QC) results for genotyping data

**1. Collection-level SNP QC**

| | CD cases | | | | Controls | |
|---|---|---|---|---|---|---|
| | KHUH | | IMPACT | | KoGES* | |
| **1. Collection-level SNP QC** | **samples (n)** | **variants (n)** | **samples (n)** | **variants (n)** | **samples (n)** | **variants (n)** |
| 1-0. initial collection | 260 | 800,476 | 657 | 797,702 | 72,294 | 465,971 |
| 1-1. call rate ≥ 97% | 260 | 757,499 | 657 | 706,640 | 72,294 | 425,245 |
| 1-2. polymorphic variants | 260 | 652,149 | 657 | 614,204 | 72,294 | 425,245 |
| 1-3. *P* for HWE < $10^{-6}$ for controls and $10^{-10}$ for cases | 260 | 651,269 | 657 | 614,204 | 72,294 | 425,108 |

**2. QC after merging three batches**

| **2. SNP QC after merging batches** | **CD cases (n)** | **Controls (n)** | **Variants (n)** |
|---|---|---|---|
| 2-1. present in all three batches | 917 | 72,294 | 388,055 |
| 2-2. P for differential call rates between batches ≥ $10^{-8}$ | 917 | 72,294 | 322,998 |
| 2-3. call rate ≥ 97% | 917 | 72,294 | 322,998 |
| 2-4. MAF ≥ 0.5% | 917 | 72,294 | 322,998 |
| 2-5. *P* for HWE < $10^{-6}$ for controls and $10^{-10}$ for cases | 917 | 72,294 | 322,995 |
| 2-6. duplicates excluded | 917 | 72,294 | 322,994 |
| **3. Sample QC** | **CD cases (n)** | **Controls (n)** | **Variants (n)** |
| 3-1. call rate ≥ 95% & no heterozygosity outliers | 915 | 72,294 | 322,994 |
| 3-2. cryptic 1st/2nd degree relatives removed | 902 | 72,179 | 322,994 |
| 3-3. samples with gender discrepancy removed | 902 | 72,179 | 322,994 |
| **4. Pre-imputation SNP QC** | **CD cases (n)** | **Controls (n)** | **Variants (n)** |
| 4-1. present in the reference panel | 902 | 72,179 | 321,885 |
| 4-2. no allele mismatches with the reference panel | 902 | 72,179 | 321,616 |

* The KoGES provided genetic data after performing the first-round QC based on the following criteria:

i) Sample exclusion criteria: call rate (<97%), heterozygosity outliers, excessive singleton, gender discrepancy, cryptic 1st degree relatives, withdrawals and blind replicates

ii) SNP exclusion criteria: low quality SNP in any experimental batches, P for HWE < $10^{-6}$, low call rate (<95%).

HWE, Hardy-Weinberg equilibrium; MAF, minor allele frequency.

**Supplementary Table 2. Genetic associations of known loci with susceptibility to Crohn's disease in Korean populations**

| | | | | Korean dataset #1 (new) | | Korean dataset #2 (Jung *et al.*) | | Meta-analysis | | | |
|---|---|---|---|---|---|---|---|---|---|---|---|
| **Variants** | **Chr:Position (hg19)** | **Nearby Genes** | **EA/NEA** | **OR (95% CI)** | ***P*** | **OR (95% CI)** | ***P*** | **EAF** | **OR (95% CI)** | $P_{meta}$ | $P_{het}$* |
| rs2640906 | 1:8029509 | *TNFRSF9,PARK7* | A/G | 0.77 (0.69-0.85) | 1.18E-06 | 0.84 (0.77-0.93) | 3.31E-04 | 0.603 | 0.81 (0.75-0.87) | 1.30E-08 | 0.218 |
| rs11249215 | 1:25297184 | *RUNX3* | A/G | 0.79 (0.71-0.88) | 2.82E-05 | 0.78 (0.72-0.86) | 9.07E-08 | 0.545 | 0.79 (0.73-0.85) | 7.76E-11 | 0.914 |
| rs12033764 | 1:67734482 | *IL23R* | T/C | 1.24 (1.11-1.38) | 1.51E-04 | 1.27 (1.16-1.39) | 1.24E-07 | 0.471 | 1.26 (1.17-1.35) | 5.03E-10 | 0.721 |
| rs3766920 | 1:154934963 | *PAGO2,SHC1* | A/G | 1.26 (1.06-1.49) | 9.41E-03 | 1.59 (1.36-1.85) | 3.09E-09 | 0.081 | 1.43 (1.27-1.61) | 4.41E-09 | 0.0551 |
| rs10196794 | 2:234192616 | *ATG16L1* | A/G | 1.30 (1.18-1.45) | 4.62E-07 | 1.27 (1.16-1.40) | 7.93E-07 | 0.306 | 1.29 (1.20-1.39) | 1.28E-11 | 0.748 |
| rs2975780 | 2:241563652 | *GPR35* | A/T | 0.69 (0.61-0.77) | 3.72E-11 | 0.73 (0.67-0.80) | 6.74E-11 | 0.686 | 0.71 (0.66-0.77) | 6.29E-19 | 0.403 |
| rs71055031 | 3:18760051 | *SATB1-AS1* | ATGT/A | 1.43 (1.27-1.61) | 1.59E-09 | NA | NA | 0.220 | 1.43 (1.27-1.62) | 4.85E-09 | NA |
| rs34762726 | 3:49689210 | *BSN* | A/G | 1.59 (1.32-1.93) | 1.59E-06 | 1.34 (1.13-1.59) | 8.81E-04 | 0.064 | 1.45 (1.27-1.66) | 4.28E-08 | 0.198 |
| rs73243351 | 4:38335067 | *LINC01258* | A/G | 1.31 (1.16-1.49) | 1.60E-05 | 1.59 (1.42-1.77) | 6.36E-17 | 0.199 | 1.46 (1.34-1.59) | 2.73E-18 | 0.0316 |
| rs55896948 | 4:123342114 | *IL2,IL21* | G/T | 1.28 (1.15-1.42) | 9.95E-06 | 1.21 (1.11-1.33) | 2.64E-05 | 0.467 | 1.24 (1.15-1.33) | 6.03E-09 | 0.478 |
| rs4957135 | 5:40508137 | *TTC33* | G/A | 1.28 (1.16-1.41) | 5.11E-07 | 1.27 (1.16-1.38) | 2.00E-07 | 0.420 | 1.27 (1.19-1.36) | 3.96E-12 | 0.869 |
| rs12654043 | 5:150226095 | *IRGM* | G/A | 1.27 (1.14-1.41) | 8.89E-06 | 1.20 (1.10-1.32) | 6.43E-05 | 0.573 | 1.23 (1.15-1.32) | 1.17E-08 | 0.49 |
| rs56167332 | 5:158827769 | *IL12B* | A/C | 1.25 (1.12-1.41) | 1.56E-04 | 1.34 (1.21-1.47) | 8.97E-09 | 0.326 | 1.30 (1.20-1.41) | 6.23E-11 | 0.441 |
| rs4997107 | 6:167403485 | *CCR6* | T/C | 1.14 (1.03-1.27) | 1.04E-02 | 1.29 (1.18-1.41) | 2.73E-08 | 0.367 | 1.22 (1.14-1.31) | 2.12E-08 | 0.0986 |
| rs4385425 | 7:50307334 | *KZF1* | G/A | 1.28 (1.15-1.41) | 1.94E-06 | 1.20 (1.10-1.31) | 8.08E-05 | 0.407 | 1.23 (1.15-1.32) | 4.10E-09 | 0.375 |
| rs56211063 | 9:117585897 | *TNFSF15,TNFSF8* | C/G | 2.06 (1.86-2.28) | 2.58E-45 | 2.12 (1.94-2.32) | 7.06E-60 | 0.351 | 2.09 (1.95-2.24) | 3.52E-95 | 0.694 |
| rs224135 | 10:64466802 | *ZNF365,ADO* | G/A | 0.84 (0.76-0.93) | 1.10E-03 | 0.76 (0.69-0.83) | 1.66E-09 | 0.381 | 0.79 (0.74-0.85) | 1.96E-10 | 0.123 |
| rs10786560 | 10:101315166 | *SLC25A28* | A/G | 1.42 (1.26-1.59) | 5.85E-09 | 1.27 (1.15-1.41) | 2.09E-06 | 0.305 | 1.33 (1.23-1.44) | 1.16E-12 | 0.196 |
| rs11195128 | 10:112186148 | *DUSP5* | T/C | 1.62 (1.42-1.85) | 7.58E-13 | 1.50 (1.33-1.69) | 1.97E-11 | 0.153 | 1.55 (1.42-1.70) | 5.54E-21 | 0.402 |
| rs3764147 | 13:44457925 | *CCDC122* | G/A | 1.37 (1.24-1.51) | 4.06E-10 | 1.15 (1.05-1.26) | 2.79E-03 | 0.348 | 1.25 (1.16-1.34) | 4.45E-10 | 0.0148 |
| rs60411253 | 14:35863871 | *NFKBIA* | T/C | 0.79 (0.70-0.88) | 4.08E-05 | 0.79 (0.71-0.87) | 5.77E-06 | 0.266 | 0.79 (0.73-0.85) | 4.42E-09 | 0.999 |
| rs4787458 | 16:28531287 | *IL27* | G/A | 1.47 (1.25-1.74) | 3.98E-06 | 1.44 (1.25-1.65) | 3.80E-07 | 0.104 | 1.45 (1.30-1.62) | 4.73E-11 | 0.819 |
| rs4103200 | 17:40507065 | *STAT3* | C/G | 0.77 (0.69-0.86) | 1.51E-06 | 0.74 (0.67-0.82) | 8.34E-10 | 0.315 | 0.76 (0.70-0.81) | 9.16E-14 | 0.585 |
| rs2427537 | 20:62376227 | *ZBTB46* | C/T | 0.72 (0.58-0.89) | 2.98E-03 | 0.59 (0.49-0.71) | 2.88E-08 | 0.915 | 0.64 (0.55-0.74) | 4.59E-09 | 0.182 |
| rs5754102 | 22:21916272 | *UBE2L3* | A/C | 1.36 (1.18-1.58) | 4.47E-05 | 1.25 (1.13-1.38) | 8.30E-06 | 0.251 | 1.29 (1.18-1.40) | 1.01E-08 | 0.368 |
| rs12628495 | 22:37307711 | *NCF4,CSF2RB* | T/C | 1.24 (1.08-1.41) | 1.62E-03 | 1.30 (1.18-1.43) | 1.41E-07 | 0.237 | 1.28 (1.18-1.39) | 5.26E-09 | 0.574 |

*P values for the heterogeneity in genetic effect sizes between two datasets.

Chr, Chromosome; EA, effect allele; NEA, non-effect allele; OR, odds ratio; CI, confidence interval; EAF, effect allele frequency.

**Supplementary Table 3. The 95% credible set of putative causal variants in the LILR gene cluster locus**

| Chromosome | Position (hg19) | rsID | REF | ALT | Posterior probability |
|---|---|---|---|---|---|
| **19** | **54723355** | **rs255774** | **A** | **G** | **0.383** |
| **19** | **54723546** | **rs255773** | **C** | **T** | **0.377** |
| 19 | 54723822 | rs2885369 | G | A | 0.078 |
| 19 | 54723823 | rs2885368 | T | C | 0.069 |
| 19 | 54723813 | rs8101337 | G | C | 0.055 |

Variants with a posterior probability > 0.1 are highlighted in bold.
REF, reference allele; ALT, alternative allele.

**Supplementary Table 4. The 95% credible set of putative causal variants in the *ZBTB38* locus**

| Chromosome | Position (hg19) | rsID | REF | ALT | Posterior probability |
|---|---|---|---|---|---|
| 3 | 141145473 | rs6802753 | T | C | 0.078 |
| 3 | 141146944 | rs78960151 | C | G | 0.062 |
| 3 | 141145216 | rs6762722 | A | G | 0.059 |
| 3 | 141145315 | rs6762826 | A | G | 0.057 |
| 3 | 141152220 | rs59075313 | C | G | 0.038 |
| 3 | 141148930 | rs9822817 | A | T | 0.036 |
| 3 | 141148231 | rs6810158 | T | C | 0.034 |
| 3 | 141151456 | rs56339671 | G | A | 0.032 |
| 3 | 141137035 | rs9825379 | G | A | 0.031 |
| 3 | 141150161 | rs143772616 | G | A | 0.031 |
| 3 | 141136915 | rs73872720 | C | A | 0.031 |
| 3 | 141139819 | rs79217331 | T | C | 0.031 |
| 3 | 141096518 | rs77753509 | G | A | 0.030 |
| 3 | 141152017 | rs9876694 | C | T | 0.028 |
| 3 | 141095242 | rs77316854 | T | A | 0.027 |
| 3 | 141140659 | rs16851412 | G | A | 0.027 |
| 3 | 141140861 | rs79928910 | G | A | 0.026 |
| 3 | 141140272 | rs74557261 | A | G | 0.026 |
| 3 | 141140534 | rs111518062 | C | A | 0.026 |
| 3 | 141148143 | rs6440007 | G | A | 0.026 |
| 3 | 141141904 | rs79474768 | T | C | 0.026 |
| 3 | 141148158 | rs115037602 | G | T | 0.026 |
| 3 | 141147819 | rs75220288 | C | A | 0.021 |
| 3 | 141118232 | rs74888405 | T | C | 0.015 |
| 3 | 141154542 | rs6440008 | T | C | 0.015 |
| 3 | 141087623 | rs1863868 | T | C | 0.013 |
| 3 | 141145402 | rs55715186 | T | C | 0.012 |
| 3 | 141143430 | rs10513137 | G | A | 0.011 |
| 3 | 141111009 | rs76983194 | A | C | 0.010 |
| 3 | 141142391 | rs6776991 | C | G | 0.010 |
| 3 | 141110287 | rs73872711 | C | G | 0.009 |
| 3 | 141138833 | rs9869102 | A | G | 0.008 |
| 3 | 141126825 | rs57345461 | A | T | 0.006 |
| 3 | 141148698 | rs9856584 | C | A | 0.006 |
| 3 | 141147414 | rs7650602 | T | C | 0.006 |
| 3 | 141093285 | rs7624084 | T | C | 0.005 |
| 3 | 141142691 | rs6440006 | G | A | 0.005 |
| 3 | 141127710 | rs58473751 | C | G | 0.005 |
| 3 | 141136727 | rs9858405 | A | C | 0.005 |

REF, reference allele; ALT, alternative allele.

**Supplementary Table 5. Lead eQTLs for the genes in the MHC class I-mediated antigen presentation pathway and interaction analysis**

| Gene symbol | Ensembl gene ID | Chromosome | Start (hg19) | End (hg19) | $P_{meta}$ < 5e-8 in our study? |
|---|---|---|---|---|---|
| *FCER1G* | ENSG00000158869 | 1 | 161185024 | 161190489 | No |
| *MR1* | ENSG00000153029 | 1 | 181002510 | 181031074 | No |
| *MFSD6* | ENSG00000151690 | 2 | 191273081 | 191373931 | No |
| *ERAP1* | ENSG00000164307 | 5 | 96096514 | 96143803 | No |
| *ERAP2* | ENSG00000164308 | 5 | 96211690 | 96255407 | No |
| *LNPEP* | ENSG00000113441 | 5 | 96271098 | 96373217 | No |
| *SAR1B* | ENSG00000152700 | 5 | 133936839 | 133984961 | No |
| *HFE* | ENSG00000010704 | 6 | 26087509 | 26098571 | Yes |
| *HLA-F* | ENSG00000204642 | 6 | 29690552 | 29706305 | Yes |
| *HLA-G* | ENSG00000204632 | 6 | 29794744 | 29798902 | Yes |
| *HLA-H* | ENSG00000206341 | 6 | 29855529 | 29858259 | Yes |
| *HLA-A* | ENSG00000206503 | 6 | 29909037 | 29917349 | Yes |
| *HLA-E* | ENSG00000204592 | 6 | 30457286 | 30461971 | Yes |
| *HLA-C* | ENSG00000204525 | 6 | 31236526 | 31239907 | Yes |
| *HLA-B* | ENSG00000234745 | 6 | 31321649 | 31334844 | Yes |
| *TAP2* | ENSG00000204267 | 6 | 32789610 | 32806516 | Yes |
| *TAP1* | ENSG00000168394 | 6 | 32812986 | 32821593 | Yes |
| *TAPBP* | ENSG00000231925 | 6 | 33267471 | 33282061 | Yes |
| *AZGP1* | ENSG00000160862 | 7 | 99564343 | 99573665 | No |
| *IKBKB* | ENSG00000104365 | 8 | 42128820 | 42189978 | No |
| *IDE* | ENSG00000119912 | 10 | 94211441 | 94333853 | No |
| *TAPBPL* | ENSG00000139192 | 12 | 6560856 | 6575683 | No |
| *CLEC4A* | ENSG00000111729 | 12 | 8276213 | 8291203 | No |
| *ABCB9* | ENSG00000150967 | 12 | 123405498 | 123466196 | No |
| *PDIA3* | ENSG00000167004 | 15 | 44038592 | 44065477 | No |
| *B2M* | ENSG00000166710 | 15 | 45003556 | 45011049 | No |
| *ACE* | ENSG00000159640 | 17 | 61554422 | 61575741 | No |
| *CALR* | ENSG00000179218 | 19 | 13049392 | 13055303 | No |
| *IFI30* | ENSG00000216490 | 19 | 18283972 | 18288927 | No |

The list of genes was extracted from the gene ontology (GO) term ANTIGEN PROCESSING AND PRESENTATION OF PEPTIDE ANTIGEN VIA MHC CLASS I (GO:0002474). Disease association of each gene was identified based on the CD association $P_{meta}$ values of the variants in the +/- 500 kb flanking regions of the gene.

**Supplementary Table 6. Lead eQTLs for genes in the MHC class I-mediated antigen presentation pathway and interaction results**

| eQTL results from Japanese individuals | | | | | | | This study | |
|---|---|---|---|---|---|---|---|---|
| eGene | Tissue | Lead eQTL | Pos (hg19) | Allele1 | Allele2 | $P_{adj}$ for eQTL | $P_{meta}$ for CD risk | *P* for interaction with rs255774 on CD risk |
| *HLA-F* | B_cells | chr6:29690668:D | 29690668 | AAAG | A | 1.87E-03 | NA | NA |
| *HLA-F* | CD8+T_cells | rs7738786 | 29685026 | A | G | 4.66E-03 | 1.37E-09 | 0.328 |
| *HLA-F* | Monocytes | rs116255195 | 29677787 | C | T | 7.21E-03 | 4.08E-02 | NA |
| *HLA-F* | NK_cells | rs7738919 | 29685025 | C | T | 5.50E-05 | 1.37E-09 | 0.328 |
| *HLA-F* | Peripheral_blood | rs113870146 | 29849479 | C | T | 1.78E-07 | 4.37E-06 | NA |
| *HLA-G* | B_cells | rs114701937 | 29956405 | G | A | 1.98E-02 | 0.497 | NA |
| *HLA-G* | Peripheral_blood | rs114701937 | 29956405 | G | A | 2.05E-03 | 0.497 | NA |
| *HLA-H* | B_cells | rs114273758 | 29825434 | A | G | 9.84E-31 | 0.388 | NA |
| *HLA-H* | CD4+T_cells | rs114720641 | 29922885 | C | T | 1.09E-30 | 0.126 | NA |
| *HLA-H* | CD8+T_cells | rs114720641 | 29922885 | C | T | 4.41E-31 | 0.126 | NA |
| *HLA-H* | Monocytes | rs115988571 | 29908892 | G | A | 1.22E-31 | 0.115 | NA |
| *HLA-H* | NK_cells | rs114691803 | 29911360 | A | G | 7.77E-30 | 0.116 | NA |
| *HLA-H* | Peripheral_blood | rs114720641 | 29922885 | C | T | 4.86E-29 | 0.126 | NA |
| *HLA-A* | Monocytes | rs112909485 | 29937740 | C | T | 1.26E-03 | 1.26E-02 | NA |
| *HLA-E* | Monocytes | rs114386415 | 30404403 | A | G | 4.61E-02 | 0.188 | NA |
| *HLA-C* | B_cells | rs148197692 | 31073093 | T | C | 2.61E-04 | 0.944 | NA |
| *HLA-C* | CD4+T_cells | rs116709955 | 31249127 | G | A | 3.71E-05 | 0.646 | NA |
| *HLA-C* | Monocytes | rs116471209 | 31202575 | C | G | 1.53E-11 | 0.637 | NA |
| *HLA-C* | NK_cells | chr6:31236715:D | 31236715 | GC | G | 4.07E-03 | 0.763 | NA |
| *HLA-C* | Peripheral_blood | rs76993737 | 31241761 | A | T | 4.82E-04 | 0.194 | NA |
| *HLA-B* | B_cells | rs116144359 | 31239869 | T | C | 2.89E-03 | 1.98E-03 | NA |
| *HLA-B* | CD4+T_cells | rs2523580 | 31328245 | A | G | 8.00E-03 | 0.763 | NA |
| *TAP2* | B_cells | rs114501627 | 32684387 | A | G | 6.27E-03 | 1.19E-03 | NA |
| ***TAP2*** | **CD4+T_cells** | **rs144541539** | **32743835** | **C** | **T** | **3.39E-08** | **9.29E-14** | **4.11E-04** |
| ***TAP2*** | **CD8+T_cells** | **rs144541539** | **32743835** | **C** | **T** | **3.41E-07** | **9.29E-14** | **4.11E-04** |
| *TAP2* | Monocytes | rs4148874 | 32797488 | C | T | 3.56E-06 | 7.30E-09 | 0.185 |
| *TAP2* | NK_cells | chr6:32796856:D | 32796856 | GAC | G | 2.23E-06 | NA | NA |
| *TAP2* | Peripheral_blood | rs114779764 | 32731710 | A | G | 1.63E-03 | 3.68E-06 | NA |
| *TAPBP* | CD8+T_cells | rs140616813 | 33021474 | C | T | 3.36E-02 | 0.351 | NA |

Lead eQTLs in six immune cell types of Japanese individuals were retrieved from http://jenger.riken.jp/en/result. *P* values for eQTLs were corrected for multiple testing using gene-wide Bonferroni correction. Epistatic interaction was assessed only when a lead eQTL has a significant main effect on CD susceptibility ($P_{meta}$<5x10$^{-8}$). Variants with significant interaction with rs255774 are highlighted in bold.

**Supplementary Table 7. CD-risk HLA alleles of the MHC class I genes and interaction results**

| HLA alleles (classical alleles or amino acid residues) | *P* for CD risk | *P* for interaction with rs255774 |
|---|---|---|
| HLA_B*15 | 3.25E-10 | 0.115 |
| HLA_B*15:01:01:01 | 6.16E-09 | 0.660 |
| HLA_B*46 | 1.63E-10 | 0.105 |
| HLA_B*46:01:01 | 3.32E-10 | 0.0973 |
| AA_B_76_31324509_exon2_E | 8.20E-10 | 0.135 |
| AA_B_76_31324509_exon2_V | 8.14E-10 | 0.135 |
| AA_B_69_31324530_exon2_AT | 8.20E-10 | 0.135 |
| AA_B_69_31324530_exon2_R | 8.14E-10 | 0.135 |
| AA_B_66_31324539_exon2_IN | 8.20E-10 | 0.135 |
| AA_B_66_31324539_exon2_K | 8.14E-10 | 0.135 |
| AA_B_66_31324539_exon2_KQ | 8.14E-10 | 0.135 |
| AA_B_66_31324539_exon2_KS | 8.14E-10 | 0.135 |

After imputing HLA variants from the newly generated GWAS dataset, we identified CD associations of classical alleles and amino acids of *HLA-B* ($P<5x10^{-8}$) but not *HLA-A* and *HLA-C*.

CD, Crohn's disease.

**Supplementary Table 8. Synergistic interaction effect of the two quantitative trait loci for *TAP2* and LILR genes on susceptibility to Crohn's disease**

| No. of CD-risk alleles of the *TAP2* lead eQTL rs9276627 | No. of cases/controls | CD association of the LILR variant rs255774 | | |
|---|---|---|---|---|
| | | EAF (cases/control) | OR (95% CI) | *P* value |
| 0 | 9/1,627 | 0.389/0.470 | 0.723 (0.276-1.891) | 0.508 |
| 1 | 177/18,396 | 0.452/0.477 | 0.904 (0.731-1.118) | 0.352 |
| 0 or 1 | 186/20,023 | 0.448/0.477 | 0.894 (0.727-1.100) | 0.288 |
| 2 | 716/52156 | 0.557/0.476 | 1.392 (1.251-1.548) | $1.11 \times 10^{-9}$ |

Effect and non-effect alleles of rs9276627 are *C* and *T*, respectively.

CD, Crohn's disease; eQTL, expression quantitative trait locus; EAF, effect allele frequency; OR, odds ratio; CI, confidence interval.

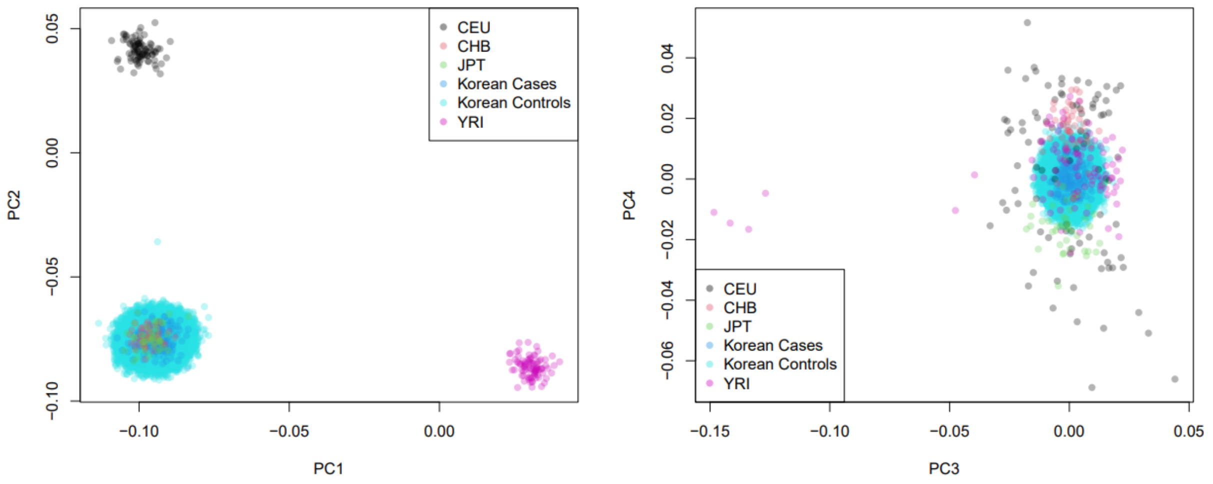


**Supplementary Figure 1. Principal component analysis results.** The top four principal components (PCs) were estimated from 902 Korean patients with CD and 72,179 healthy controls by projecting the genetic data on PC axes estimated from 90 European (CEU), 90 African (YRI), 45 Chinese (CHB), and 45 Japanese (JPT) individuals of the International HapMap Project phase 3 (release 23).
